# Understanding Data Differences across the ENACT Federated Research Network

**DOI:** 10.1101/2025.01.17.25320686

**Authors:** Taowei D Wang, Darren W Henderson, Griffin M Weber, Michele Morris, Eugene M Sadhu, Shawn N Murphy, Shyam Visweswaran, Jeff G Klann

**Affiliations:** Department of Neurology, Massachusetts General Hospital, Boston, MA 02114, USA; Center for Clinical and Translational Science, University of Kentucky, Lexington, KY 40536, USA; Department of Biomedical Informatics, Harvard Medical School, Boston, MA 02115, USA; Department of Biomedical Informatics, University of Pittsburgh, Pittsburgh, PA 15206, USA; Laboratory of Computer Science, Massachusetts General Hospital, Boston, MA 02114, USA

## Abstract

**Objective:** Federated research networks, like Evolve to Next-Gen Accrual of patients to Clinical Trials (ENACT), aim to facilitate medical research by exchanging electronic health record (EHR) data. However, poor data quality can hinder this goal. While networks typically set guidelines and standards to address this problem, we developed an organically evolving, data-centric method using patient counts to identify data quality issues, applicable even to sites not yet in the network.

**Materials and Methods:** We distribute high-performance patient counting scripts as part of Integrating Biology at the Bedside (i2b2), which all ENACT sites operate. They produce counts of patients associated with ENACT ontology terms for each site. At the ENACT Hub, our pipeline aggregates site-contributed counts to produce network statistics, which our self-service web application, Data Quality Explorer (DQE), ingests to help sites conduct data quality investigation relative to the network.

**Results:** Thirteen ENACT sites have contributed their patient counts, and currently seven sites have signed up to use DQE to analyze data quality issues. We announced a call to all ENACT sites to contribute additional patient counts.

**Discussion:** Identifying site data quality problems relative to the network is novel. Using a metric based on evolving network statistics complements rigid data quality checks. It is adaptable to any network and has low barriers of entry, with patient counting being the sole requirement.

**Conclusion:** We implemented a metric for conducting data quality investigation in ENACT using patient counting and network statistics. Our end-to-end pipeline is privacy-preserving and the underlying design is generalizable.

## INTRODUCTION

Clinical research data networks promise rapid discovery and feedback into the healthcare system. However, electronic health record (EHR) data is fraught with problems due to eccentricities in data collection, organization/structure, and errors in data processing (e.g., during Extract Transform Load, or ETL). Federated data networks introduce further room for quality problems - non-uniformity of data across sites and the need for data harmonization across locations using different terminologies and coding practices. The quality of data in a research network can directly impact the quality of research that relies on it, even at early stages of study design and planning. To effectively convert data to knowledge, methods must be developed to help researchers understand how the data are arranged in these different settings.

The National Institutes of Health’s (NIH) National Center for Advancing Translational Sciences (NCATS) program funds a large federated clinical research data network called Evolve to Next-Gen Accrual of patients to Clinical Trials (ENACT).<u>[1,2]</u> The network consists of local EHR data repositories that use either the Informatics for Integrating Biology at the Bedside (i2b2) or the Observational Medical Outcomes Partnership (OMOP) common data models.<u>[3,4]</u> These are linked via the Shared Health Research Information Network (SHRINE) software platform.<u>[5,6]</u> The ENACT network consists of 52 participating sites across the United States, covering over 140 million patients. Queries are “real-time” in that once investigators specify their search criteria, sites immediately begin returning (obfuscated) counts of the number of matching patients. The patient counts enable investigators to assess feasibility of any planned study and to identify potential collaborating sites.

The i2b2 platform is used at over 250 locations worldwide, including in large data research networks like the National Patient-Centered Clinical Research Network (PCORnet) and ENACT.<u>[7–9]</u> OHDSI reports over 500 databases using the OMOP data model worldwide.<u>[10]</u>

Key to i2b2’s design is its “ontology” system, which supports hierarchies to encode the granular knowledge of biomedicine. ENACT similarly maintains a set of standard ontologies to which each site maps its own local ontologies. This local-to-standard ontology mapping enables the network sites to harmonize the types of data that are shared. The ontology design also allows OMOP to be queried; ENACT distributes a version of their ontology that queries OMOP tables in which i2b2-style standard codes are replaced with OMOP identifiers.<u>[11,12]</u>

Despite standardization with ontologies, a great deal of variation across sites exists. Each local site must map their local data to the standard ontologies. Mapping local data to standard ontologies is nuanced and error-prone for several reasons: (1) Strategies for mapping data differ based on local infrastructure, policies, and processes. For example, some sites map to standard codes during ETL, whereas others utilize the ontology system to dynamically manage mappings.<u>[13]</u> (2) Many codes captured during clinical practice are transformed by professional coders to satisfy reimbursement or reporting requirements. (3) Standard terminologies are often not assigned at the source, leaving informatics teams to encode facts using their best educated guess. (4) Modern ontologies contain hundreds of thousands of terms (ENACT ontology has over 800,000), and sites tend to use only a small portion, so a laboratory test like hemoglobin A1C could be encoded with any of several valid LOINC codes.

Other clinical data networks have tried to surmount the data quality problems with their own tools. OHDSI provides the data characterization tool Automated Characterization of Health Information at Large-scale Longitudinal Evidence Systems (ACHILLES).<u>[14]</u> More recently, the Data Quality Dashboard (DQD) provides a snapshot view of common data quality issues through 24 data check types that check Plausibility, Conformance, and Completeness.<u>[15,16]</u> PCORnet offers comprehensive data characterization reports with a similar combination of conformance checks and data characterization graphs.<u>[17]</u>

The ENACT network, built on i2b2, creates a unique opportunity for a flexible data quality methodology built around its unique star-schema data structure. Rather than delineating fact types through table definitions (e.g., CONDITION, DRUG, LAB), the i2b2 ontology system uniquely defines the possible facts in the data. Facts are stored as generic objects in a single fact table, with ancillary dimension tables providing additional context about the facts. Rather than building hundreds of specific data quality checks, ENACT gives us the opportunity to engage in a measurement of quality through the ontology and relative to the network. The absence of a set of codes might not indicate a quality issue (perhaps the codes are esoteric and seldom used), but if every other site in the network has data with these codes, it is a strong indicator that the missing data are a quality issue.

In ENACT, we could envision analyzing the distribution and study similarities/differences in ontology usage at sites when i2b2 sites are linked in data networks. This could be accomplished by counting the number of patients with each ontology term (and all its sub-terms). Analyzing such counts and comparing them across sites would enable us to answer questions about missingness, mapping variation, and outliers and averages.

Ontology usage in the data network could then be paired with a self-service tool to empower sites to explore, identify, and focus on potential data quality issues relative to the network. We employ techniques to reduce users’ cognitive load and enable visual pattern search in large ENACT ontologies.<u>[18]</u> We opt for the classical node-link tree visualization to best convey ontological structures, like many recent ontology visualization efforts, instead of other well-known alternatives.<u>[19–22]</u> Utilizing network ontology usage for the purpose of data quality checks is novel, and it offers several benefits over the standard-based data quality methods. It enables sites to compare its numbers against the network in a mutually privacy-preserving way: site data remain private and network statistics are aggregated. As the network grows, network statistics can become more reliable and trusted by virtue of large sample sizes. Ontology usage across the network is immediately apparent to site users. ENACT can track ontology usage changes over time to find interesting trends or responses to major health events such as the COVID-19 pandemic.

## OBJECTIVE

We propose and deploy an approach for gathering, aggregating, and then visualizing patient-count data in an intuitive tool for data quality. First, we develop and deploy high-performance patient counting tools across the network. Next, we introduce the Data Quality Explorer (DQE), a self-service, interactive visualization web tool, to enable sites -- even those not yet contributing patient count data to the ENACT network -- to detect and identify potential data quality issues such as unexplained outliers or missing data. Any site that has run the patient counting tool can use the DQE to compare their data to the current network statistics.

## MATERIALS AND METHODS

We created a workflow to collect patient counts from sites, compute network statistics, and conduct data quality checks privately. To actualize this, we developed several tools that work in unison.

### High Performance Patient Counting

The core strategy for ENACT data quality relies on utilizing the total number of patients with data for each concept in the database. i2b2 has for many years had a database field called “totalnum” in the ontology tables to store a number representing counts of patients. Its original purpose was for server-side query optimization. i2b2 exposes these “patient counts” in its web interface, allowing users at individual i2b2 sites to see the single-item counts in the ontology tree. This allows users to estimate the maximum population size when building cohort queries. The i2b2 interface does not offer a mechanism to see this information in SHRINE networks, in part due to privacy concerns by sites about exposing patient counts.

A mechanism to efficiently count the number of patients for each concept has evolved more recently. This is challenging because a typical i2b2 instance has an ontology of a half-million elements and a fact table with hundreds of millions of rows. We have implemented this as SQL scripts that can interact directly with the database, leveraging the set operations and indexing that make relational databases so efficient. Careful attention was paid to optimizing temporary computation space and converting data types to more space-efficient versions.

The method is, for each element of the hierarchy:

1. Collect all associated codes.

a. For fact-based values, this involves recursing down the tree and gathering all the child codes. The node for ICD-9 250 would also include all codes in the range 250.01-259.99.
b. For dimension-based values, this is a pre-computed set of codes listed in the ontology (e.g., ‘Race:White’ could be associated with codes ‘w’,’WHITE’,’caucasian’,’non-asian/non-black’)
2. Count all patients with one of these codes in the target table.

a. For fact-based values, this is usually the observation_fact table. However, i2b2 now supports multi-fact tables, in which a different table name can be specified in the ontology, allowing counts on “fact tables” in different data models (e.g., ‘drug_exposure’ in OMOP).
b. For dimension-based values, this is usually the patient_dimension or visit_dimension.

i2b2 1.8.0 (released 2023) introduced an optimized version of this “totalnum” script for SQL Server that uses a transitive closure table. A transitive closure is a long table that lists ancestor-descendent pairs. This essentially pre-computes the collection of child codes (step 1a above) for every node in the ontology. This allows the counting to be calculated with a simple SQL join operation, which tremendously speeds the process, often reducing compute time by 10x.

The SHRINE network client obfuscates counts by the following algorithm:

1. Generate some gaussian noise with standard deviation of 6.5
2. If the absolute value of the noise is greater than 10, use 10
3. Round the result to the nearest 5
4. Add the noise to the rounded result
5. If the rounded result is less than 10 then return “<10”, otherwise return the rounded result.

We implement a version of this algorithm that can compute these obfuscated counts within the database and store the results in a “totalnum_report” table along with the item key, which is a consistent identifier across sites in a SHRINE network. Administrators can export these files and optionally contribute them to ENACT for inclusion in the network-wide methodologies described below.

To support network-oriented data quality, we developed an aggregation pipeline that collects the reports and automatically calculates statistics on the collected data. This pipeline produces a network statistics file for each of the ENACT domain ontologies. Each file contains statistics for all ontology items in that ontology. Patient counts are never exposed in this file but are always converted to a percentage. It is calculated by dividing the (obfuscated) number of patients having the condition described by the ontology item by the (obfuscated) number of total patients at the site. We then use the sites’ percentages to derive the following network-wide descriptive statistics: mean, standard deviation, quartiles (1st quartile, median, 3rd quartile), min, and max. These measures of central tendency and variability can help any site with a totalnum_report detect possible data quality problems.

### Exploring Data for Quality Issues

The ENACT Data Quality Explorer (DQE) supports data quality efforts at sites. We identified two major features that would help with common data quality issues based on patient counts:

- **Outlier detection:** The overall percentage of patients with specific facts will generally be similar across hospitals, unless there is a known difference in demographics (e.g., the hospital is a tertiary care center that specializes in rare neurological diseases). Therefore, fact types at sites that are much less/more frequent than the average could point to possible ETL problems.
- **Missing data detection:** No individual site will have data with all 800k fact types.

Rather, local coding conventions cause different specific codes to be used at sites. For example, sites will use different LOINC codes for lab collections (LOINC codes are very granular and specify both what was collected and how it was collected). By taking advantage of i2b2’s hierarchical terminology arrangement, we can provide tools that highlight clinically cohesive groups to help identify which data is meaningfully missing. Of special importance is when a high-level term in the hierarchy is completely missing (e.g., no patients have diabetes), as this likely points to an ETL error.

Design of DQE follows the information-seeking mantra “*Overview first, zoom and filter, then details on demand.*”<u>[23]</u> A horizontal version of the *Reingold–Tilford “tidy” algorithm* is used to layout the overview tree. Its color and size-encodings help users to detect outliers and missing data. To visualize the tree more efficiently, DQE builds on the *SumTree* technique to enhance the tree view by displaying summary statistics relevant to data quality.<u>[24]</u> Finally, details of every concept are available on demand, and can be filtered using dynamic query techniques.<u>[25]</u>

## RESULTS

### Data Quality Workflow

We implemented the workflow shown in Figure 1. The patient counting scripts for the three database platforms – SQL Server, PostgreSQL, and Oracle – are distributed with i2b2 1.8.0+ (a requirement for ENACT sites).<u>[26]</u> We recommend that site administrators run these scripts periodically (e.g., after each major data refresh).

**Figure 1.**
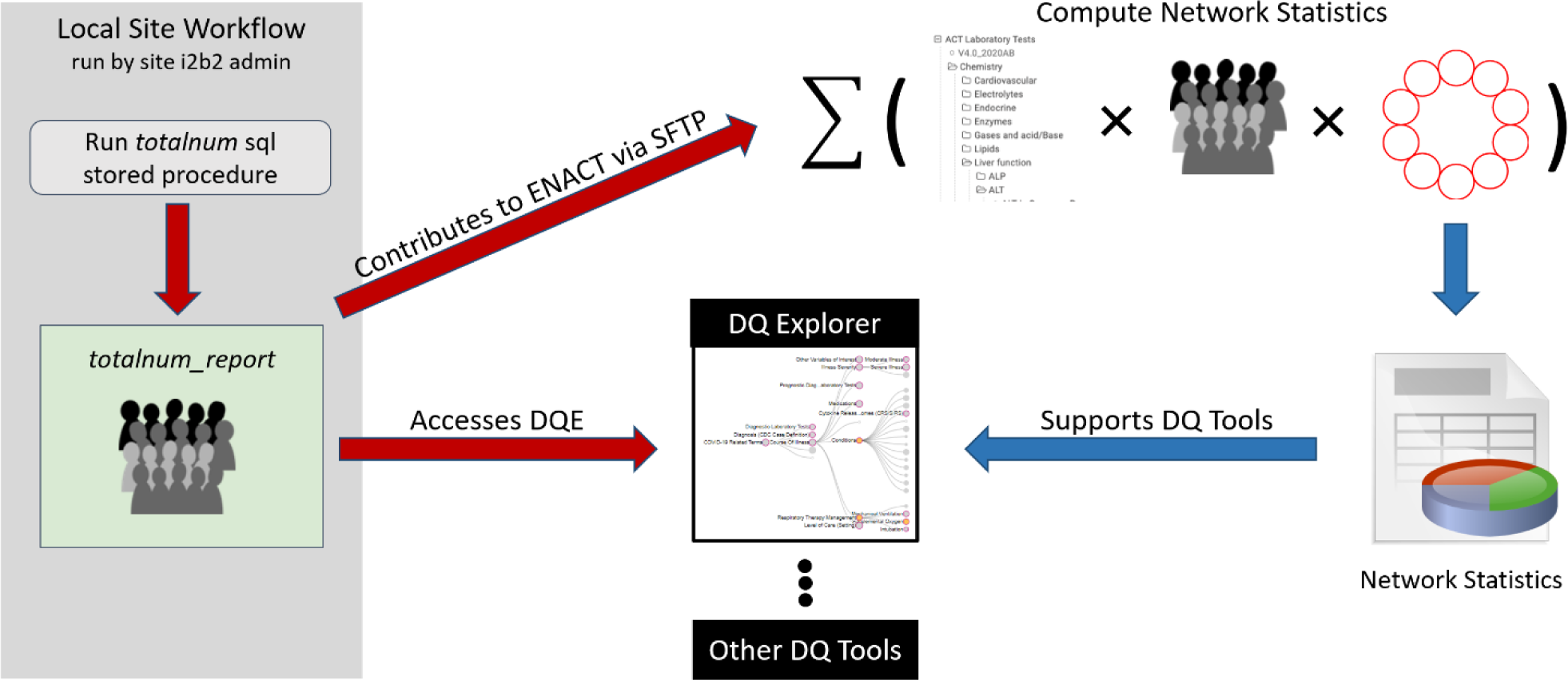
Red arrows indicate the workflow taken by ENACT site administrator. Blue arrows indicate the ENACT SHRINE Hub’s computation and dissemination of network statistics, which the Data Quality Explorer (DQE) consumes.

Sites are encouraged to export these distinct patient-per-concept counts to a file and contribute back to ENACT for inclusion in the network statistics. This is done through Secure File Transfer protocol (SFTP) to the ENACT SHRINE Hub. From here, a Hub administrator downloads the counts and executes scripts to compute the following descriptive statistics: mean, standard deviation, quartiles (1st quartile, median, 3rd quartile), min, and max. These are centrally updated as sites contribute their latest counts.

### ENACT Data Quality Explorer (DQE)

The centerpiece of this workflow is the Data Quality Explorer (DQE), a self-service, interactive visualization web tool. The DQE offers users three perspectives (traditional tree, sumTree, and data table) to inspect their data. The tool empowers sites -- even those not yet contributing their totalnum reports to the ENACT network -- to detect and identify potential data quality issues. Any site that has executed the patient counting totalnum script can use DQE to compare its own data against the current network statistics. DQE is centrally hosted on the ENACT network website<u>.[27]</u> It is a web application based on HTML and javascript. It uses the D3 library for visualization, rendering, databinding, and animation. It uses the JSZip library for decompression. It also uses JQuery, split.js, and DataTables libraries for user interface.

When DQE loads a site’s totalnum, the data remains in the browser and is never exposed. Users can select one of eleven ENACT domain ontologies to visualize and explore their data relative to the network using the three views described below.

**(Traditional Tree View)** The ontology is displayed as a classical node-link tree (Figure 2). Circles represent concepts. Lines represent subsumption relationships. The root concept is the left-most node. Its descendants are laid out to its right. Circle sizes encode patient counts. Concepts with no counts have no fill.

**Figure 2.**
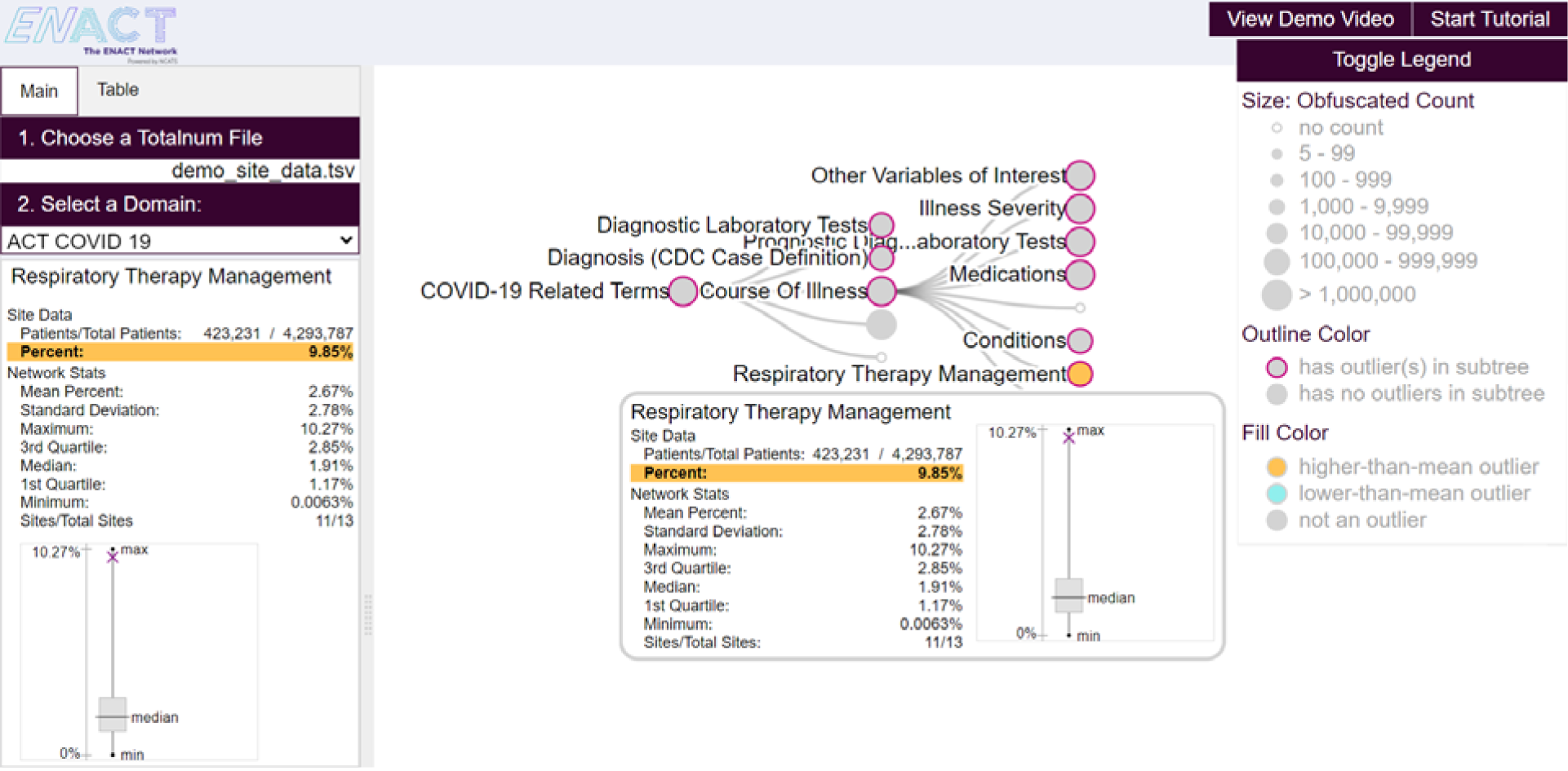
DQE’s control panel (left) allows users to load their site data and select an ENACT domain ontology to view. In this case, the ACT COVID-19 ontology is loaded. The default traditional tree view (right) enables interactive exploration of the ontology. Purple-outlined nodes indicate existence of outliers in their subtrees, and orange color signify higher-than-network mean outliers. Detailed data for the outlier Respiratory Therapy Management is shown both in the popup and in the panel to the left. Site data, network statistics, and a boxplot describing how the site data compares with the centrality and spread of network data. A legend on the right edge displays various size/color encodings.

Every concept’s site percentage is compared with its corresponding network statistics. A concept is considered an outlier if its site percentage is two-standard-deviations away from network’s mean. Higher-than-network-mean and lower-than-network-mean outliers are highlighted in orange and blue respectively. All ancestors of outliers have purple outlines for users to discover outliers.

Mousing over a circle reveals detailed data in a popup. The popup includes a table consisting of the site data, network statistics, and the count of sites contributing to the network statistics. A site count tells users whether the network statistics for that concept are reliable. The popup includes a boxplot showing the spread of the network statistics and the site percentage. The same information is displayed in a fixed panel on the left so detailed data can be captured for investigating data quality issues beyond DQE.

**(SumTree View)** A summary tree view (SumTree) is an extension to the traditional tree view and an alternative way to explore outliers by inspecting outlier distributions grouped by height. It is a linked list where each node represents a subtree height. Counts of outliers at all heights are immediately visible. In Figure 3 (1), The subtree of ***Conditions*** has height of 4, and therefore its SumTree has 4 nodes (SumNodes). Each SumNode shows the ratio of outlier counts over the total number of concepts at the same depth. Clicking on a concept toggles the traditional tree view and the SumTree view. Multiple SumTrees can be shown simultaneously.

**Figure 3.**
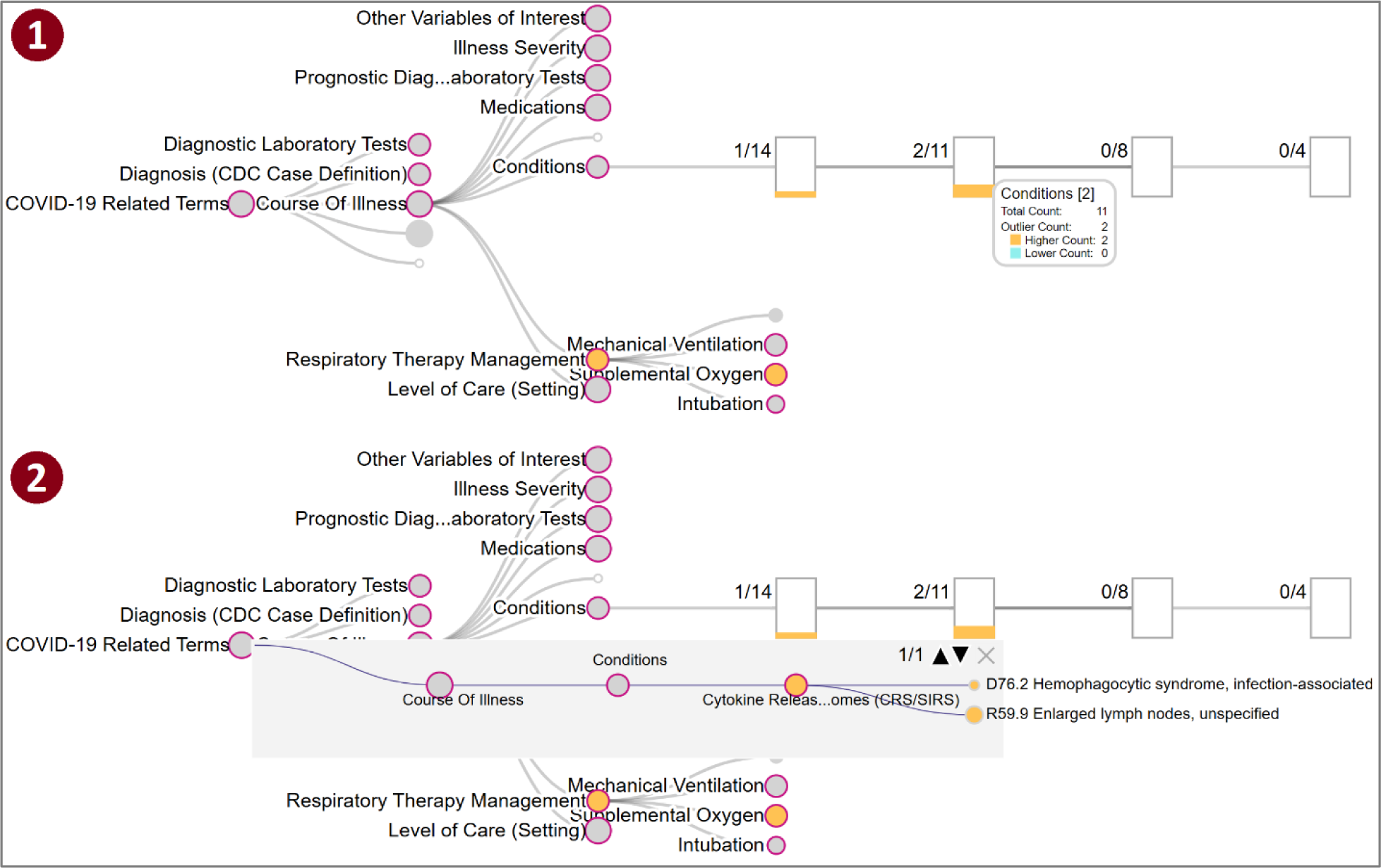
The SumTree view shows the 4-levels subtree rooted at Conditions in (1). Ratios of outliers over all nodes at each level are shown. Highlighting a rectangular node (SumNode) shows the details in a popup. Clicking on a SumNode reveals its outliers and their paths from the root in a gray inset. For example, (2) shows the overlaying inset of the 2 outliers, which share the same parent, for the SumNode labeled “2/11”.

Using the SumTree view, users can focus on identifying clusters of outliers, which may suggest data quality issues. Clicking on a SumNode shows outlier details in an inset (Figure 3 (2)). In the inset, outliers are grouped by their parents so all sibling outliers shown together. The inset displays the paths from the root to all outliers in a parent group. Users can cycle through parent groups to inspect all outliers. This view reveals whether ancestors of an outlier are also outliers themselves. If so, users may choose to focus on the outlier ancestors as they may have an outsized influence on its descendants being outliers.

**(Table View)** The table view provides a more surgical approach to outliers. It consists of a table where each row is a concept, and each column is an attribute; a set of filter controls; and a connection to visualization (4). There are seven columns in the table: *name*, *count*, *percent*, *depth* (in the ontology tree), *>2stdv* (whether an outlier), *>max* (whether site percent is greater than the network maximum), and <min (whether site percent is less than the network minimum). Concepts can be column-sorted in either direction.

The dynamic filters enable users to specify the criteria of concepts they want. Every table column has a corresponding filter control. For example, a text field for the *name* column allows users to find concepts whose name *contains* the input text. For numeric values (i.e. *count*, *percentage, depth*), users can set a range of values. Categorical data columns (i.e. *>2stdv*, *>max*, *<min*) have checkboxes, allowing users to select only the values they are interested in. Multiple column filters apply concurrently. The resulting table contains only concepts that satisfy all filters. All filters are dynamic; filter changes immediately update the table to provide immediate feedback. Clicking on a table row triggers DQE to expand the path from root to the target concept in a controlled animation, showing the concept’s location, ancestors, and siblings.

### Data Quality Explorer Use Case

Figures 4 and 5 illustrate an exploratory investigation of higher-than-network-mean outliers in the ontology ***ACT Diagnoses ICD10*** (\ACT\Diagnosis\ICD10\V2_2018AA\A20098492\) using DQE. Users first search for extreme outliers by setting the filters as shown in Figure 4, finding 1847 concepts. They are then ranked by the highest percentage, revealing that many of the top extreme outliers are related to malignant neoplasm of skin.

**Figure 4.**
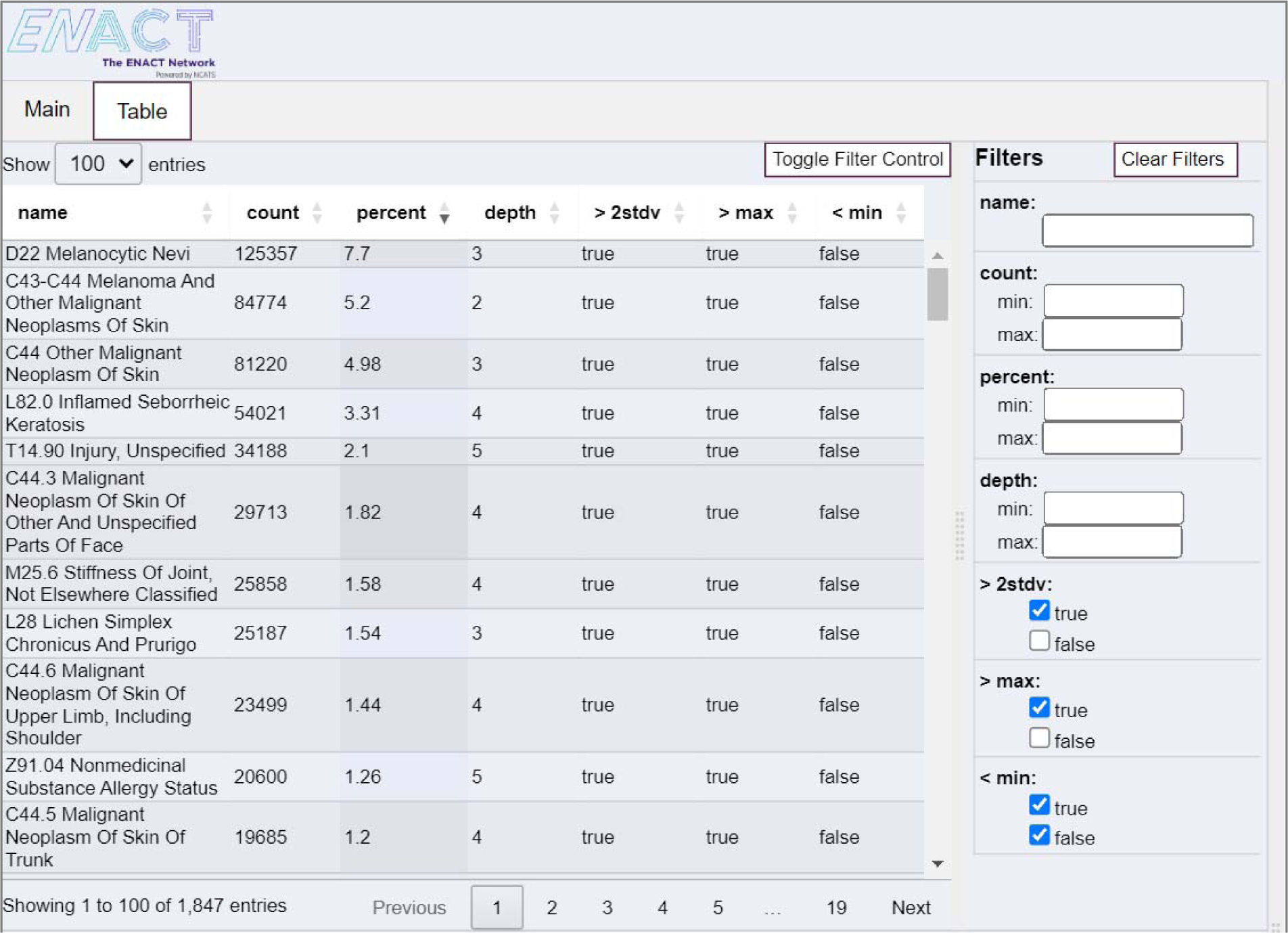
By first filtering for extreme outliers (setting >2stdv to true and >max to true) and then ranking by percent, the table produces a list of concepts where most are related to malignant neoplasm of skin. This suggests a possible cluster of extreme outliers and warrants further investigating.

Users invoke the SumTree at the root to investigate how these concepts relate to one another. In Figure 5 (1) the SumTree shows that there are 7 out of the 293 outliers in the 3rd level. ***C43-C44 Melanoma and other Malignant Neoplasm of Skin***, the extreme outlier with the second highest percentage, is one of them. Looking further in level 4, users find ***C44 Other Malignant Neoplasm of Skin***, a child of ***C43-C44***, has the third highest percentage. Users find outliers ***C44.5*** (11^th^ rank) and ***C44.52*** in levels 5 and 6 respectively in Figure 5 (2). All these descendants of ***C43-44*** collectively point to a higher-than-network mean occurrence of skin cancer. Clusters of outliers like this would often point to errors in the mapping process. However, considering this site’s subtropical climate, year-round sun, and enticing beaches, this result is perhaps unsurprising. While this specific analysis does not yield evidence of missing data or mapping problems, it illustrates the usefulness of outlier-detection techniques based on network statistics. Sites can focus on deviations that do not have explainable causes.

**Figure 5.**
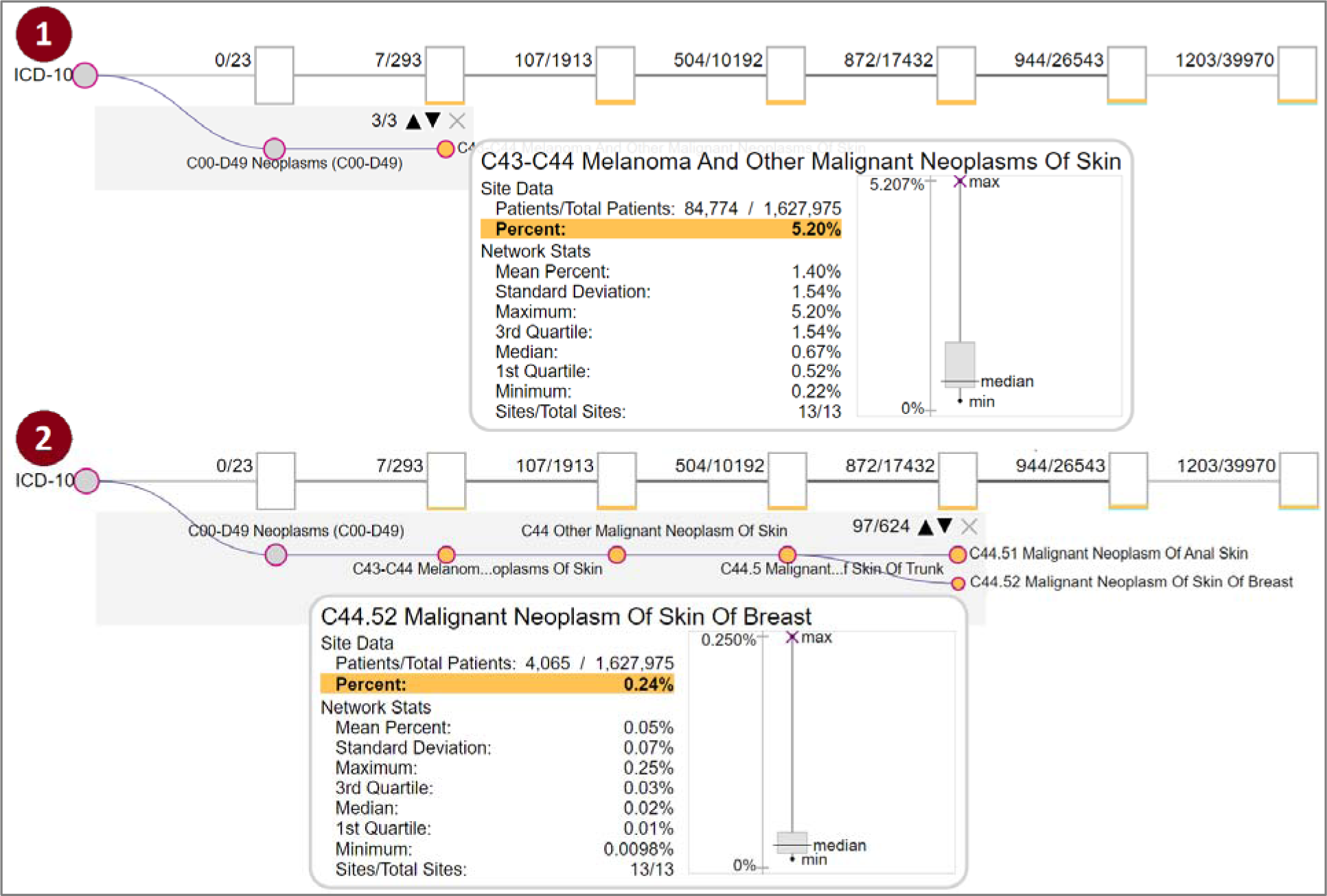
Using the SumTree, users can discover a cluster of skin cancer related outliers in the C43-C44 subtree. Many of them are the extreme outliers shown in Figure 4.

### Dissemination

We engaged in a concerted collection of patient counts in 2023. Sites were requested to run the counting script, export that data to a file, and submit it through an SFTP account to the ENACT Data Quality team. At the end of 2023, 13 ENACT sites had contributed at least one set of counts. In May 2024, we began making the DQE tool available to ENACT administrators.<u>[27]</u> As of this writing, seven sites’ data managers have DQE accounts and have been asked to give us feedback on their experiences. At present, we are soliciting a new round of patient count submissions, which will power updated network statistics and additional features (see Discussion). To date, eight sites have submitted updated counts, including two that had not previously contributed.

## DISCUSSION

Data quality in federated data networks is a complex issue. Identification of quality issues frequently requires local site-level expertise (to e.g., sort out quality issues from expected differences in populations and treatment patterns). For this reason, many data networks and data platforms have developed their own tools to help local site administrators find the issues.

Our work is unique in that from its core it is about data quality relative to the network. Rather than specific conformance checks, it enables administrators to compare their site to the network as a whole. This shifts the focus from “what data types are missing” to “in what missing data types is my site unusual”? Missing codes relative to the network can be a strong indicator of a quality issue. In other words, it enables sites to distinguish *unexpected* missing data from *expected* missing data and focus efforts on true data quality problems that can potentially be addressed.

This is particularly suited to an i2b2 network, which is defined by a large, predetermined, and shared catalog of possible fact types. The 800,000-element ENACT ontology enables a comprehensive resource of patient counts that drive an exploratory, interactive tool not unlike the i2b2 cohort query builder. The Data Quality Explorer is a specialized case of an ontology browser applied to data quality. It puts the site administrator in the driver’s seat, using their knowledge of the local data to drive the search for data quality issues. It is built on a high-performance patient-concept counting method, designed entirely in SQL, leveraging the efficient set operations and indexing of relational databases. Our approach is designed around patient privacy protection at every level. The counting tool adds Gaussian noise and a low-patient threshold to the counts, following the same design as the SHRINE network tool. The DQE folds individual site data into aggregate statistics that no longer represent any single site or patient. Finally, sites can use the tool whether they contribute to the network statistics.

Our current collection of 13 participating sites in the ENACT network and seven pilot-site-users of DQE marks a significant milestone in ENACT’s broader data quality initiative. This early success highlights the potential for scaling data quality efforts across the network. We are planning a second release of network statistics in early 2025. We also have plans in place to incorporate data quality information into the SHRINE user interface. The SHRINE tool allows ontology items to be annotated with a note through an info button that appears next to each ontology item. We plan to utilize this feature to include various network statistics. As a first pass, we will include the count of sites contributing patients to this concept. This will allow researchers to more quickly determine which query terms might lead to collaborations.

Although our DQE tool and count-collection pipeline represent a major milestone in ENACT data quality, our tooling will continue to evolve. Our second refresh of patient counts will allow us to explore a related component of data quality: tracking temporal trends. We are building visualizations for site administrators to verify that their total data is increasing over time (decreases due to ETL errors are quite common). We are also planning additional features for DQE. We plan to implement guided exploration tools - i.e. pre-compute suspected data quality issues for a site based on network statistics. Also, collecting and integrating user feedback will enable refinement of the system.

## CONCLUSION

The successful design and implementation of DQE across many sites within the ENACT network demonstrates the feasibility and value of an integrated data quality framework in large-scale health research networks. The ability to assess and improve data quality at both individual sites and across the network will enhance the reliability of research findings and foster greater confidence in the use of distributed data for clinical studies.

## AUTHOR CONTRIBUTIONS

All authors participated in data collection and the design and execution of the study. TW, JK, DH, GW contributed to the development of software. TW and JK drafted the manuscript and all authors reviewed and edited it. TW, JK, and ES managed data dissemination and data/tool hosting. MM contributed to iterative designs of software. GW contributed guidance and expertise from his own research on similar topics. SNM and SV provided overall support, guidance, and supervision. SNM, SV, and JK acquired funding. JK convened the study group, formulated the core ideas of the study, and guided the methodological design and writing.

## Data Availability

All data produced are available to ENACT members who can request a Data Quality Explorer (DQE) account online at https://enact-network.org/tools/data-quality-explorer/

## ACKNOWLEDGMENTS

We would like to thank participants in the ENACT Data Quality Work Group for their valuable feedback. We would also like to thank ENACT sites that have submitted their counts to make this work possible.

## DATA AVAILABILITY STATEMENT

Data is available to ENACT members who can request a Data Quality Explorer (DQE) account.

